# Comprehensive Analysis of Cell-Free DNA Fragmentation Across Cancer Stages

**DOI:** 10.1101/2023.11.07.23298181

**Authors:** Xin Guo, Lijuan Nie, Junjun Yan, Tinsheng Ling, Fei Zhang, Yi Chen, Mengyu Li, Wenqi Zeng, Yugen Chen, Wen-Ming Chu, Olivia Ge, Qing Guo, Dongliang Ge, Xiqiao Zhou

## Abstract

Circulating cell-free DNA (cfDNA) in the bloodstream displays cancer-derived fragmentation patterns, offering a non-invasive diagnostic avenue for cancer patients. However, the association between cfDNA fragmentation patterns and cancer progression remains largely unexplored. In this study, we analyzed this relationship using 214 whole-genome cfDNA samples across seven solid cancer types and revealed that the relation between cfDNA fragmentation patterns and cancer stages vary across cancer types. Among them, cfDNA fragmentation patterns in colorectal cancer (CRC) showed a strong correlation with cancer stages. This finding is further validated with an independent targeted cfDNA dataset from 29 CRC samples. Inspired by these findings, we designed “frag2stage”, a machine learning model that exploits cfDNA fragmentation data to differentiate cancer stages of CRC. Evaluated on two independent cfDNA datasets, our model can distinguish cancer stages of CRC with the area under the curve (AUC) values ranging from 0.68 to 0.99. The results suggest that cfDNA fragmentation patterns might carry yet undiscovered genetic and epigenetic signals, highlighting their promising potential for broader diagnostic applications in oncology.

## Introduction

Cancer staging is a major determinant of disease prognosis and a prerequisite activity in starting the process of the appropriate treatment and clinical management [1]. The tumor-node-metastasis (TNM) classification, esteemed for its consistency, simplicity, and clinically proven correlations with treatment outcomes, has become an indispensable tool in prognosticating solid tumors [2–4]. However, determining the cancer stage often requires invasive procedures, such as needle biopsies or surgeries, and in some cases a biopsy may not even be accessible. To resolve these technical and logistic difficulties, alternative non-invasive approaches are highly desired.

In recent decades, circulating cell-free DNA (cfDNA) technology has emerged as a groundbreaking non-invasive tool to access the molecular landscape of a malignancy, enabling a variety of diagnostics applications in oncology, such as early cancer detection, tumor profiling, treatment response monitoring and disease progression surveillance [5–16]. Recently, growing evidence has demonstrated that the cfDNA fragments derived from tumor cells (circulating tumor DNA or ctDNA) and the cfDNA from healthy cells are different in size. In healthy cells, the DNA wrapped around nucleosomes (∼147bp) is more protected from nucleases than the linker DNA (∼20bp) between them [17–19], which leads to a typical cfDNA size distribution peaking at ∼167bp, corresponding to the length of DNA wrapped around a nucleosome [20]. In cancer patients, the cfDNA fragment length can be altered, possibly owing to the modifications in chromatin structure caused by biological mechanisms of cancer cells [21–25]. Although contradictory analyses have been reported, the consensus of recent studies concludes the fragment size of ctDNA is overall shorter than the healthy cfDNA within the fragment region of mononucleosome (90bp to 220bp), with a mode of distribution at 145bp [22–24, 26]. Many studies have leveraged this finding using cfDNA fragment size selection techniques to improve the concentration of ctDNA [23, 27–28]. In silico, Mouliere *et al.* surveyed the ctDNA fragment size in 344 plasma samples and reported that the cfDNA in fragment sizes between 90-150bp are enriched with the mutant ctDNA [27]. Cristiano *et al.* [29] introduced DELFI, a machine-learning approach that incorporates cfDNA fragmentation profiles calculated from the whole-genome sequencing (WGS), to discriminate cancer patients from healthy individuals.

Despite these advancements, how the cfDNA fragmentation patterns evolve in relation to cancer progression remains unclear. In this study, we aim to bridge this knowledge gap by investigating the relationship between cfDNA fragmentation profiles and cancer TNM stages. As illustrated in Figure 1, we first analyzed 214 whole-genome cfDNA fragmentation profiles from seven cancer types and discovered a statistically significant correlation between the cfDNA fragmentation profiles and cancer stages in colorectal cancer. We further confirmed this association in another independent cfDNA targeted panel data from 29 CRC patients. Based upon this finding, we proposed “frag2stage”, a machine-learning model that utilizes cfDNA fragmentation profiles to distinguish CRC cancer stages.

**Figure 1.**
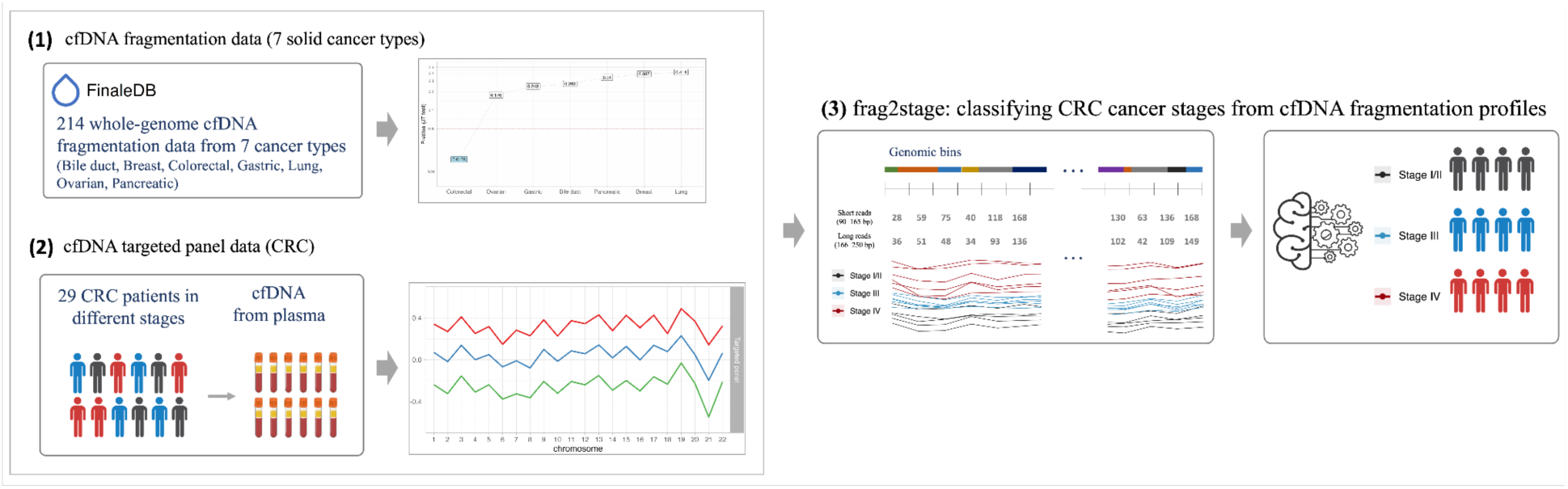
Overview of this study. (1) Association analysis of cfDNA fragmentation profiles and cancer stages: we retrieved 214 whole-genome cfDNA fragmentation data from the *FinaleDB* database [36], spanning 7 cancer types: bile duct, breast, colorectal, gastric, lung, ovarian and pancreatic cancers. The analysis indicated there exists a statistically significant association between cfDNA fragment profiles and cancer stages in colorectal cancer (CRC). (2) Further, we sequenced the blood samples from 29 colorectal cancer patients using a comprehensive cancer panel and confirmed this association in this dataset. (3) We developed “frag2stage”, a machine learning model to classify CRC cancer stages from the cfDNA fragmentation profiles of these datasets.

## Methods

### CfDNA fragmentation data collection

#### Whole-genome cfDNA fragmentation data

The whole-genome cfDNA fragmentation data, consisting of 214 cancer sequencing data spanning seven cancer types, were obtained from the FinaleDB database [36] (http://finaledb.research.cchmc.org/). We also retrieved the cfDNA fragmentation data of 242 healthy subjects as controls. This whole-genome sequencing (WGS) cfDNA data, originally published in Cristiano *et al.*, had DNA sequence composition information removed to maintain the anonymity of sensitive genotype information and subsequently archived in the *FinaleDB* database. Detailed clinical information of these patients can be found in Table 1.

**Table 1:** summary of whole-genome cfDNA fragmentation data.

|  | Stage I | Stage II | Stage III | Stage IV | Total |
| --- | --- | --- | --- | --- | --- |
| Bile Duct Cancer | 3 | 19 | 0 | 3 | 25 |
| Breast Cancer | 5 | 36 | 13 | 0 | 54 |
| Colorectal Cancer | 7 | 7 | 5 | 8 | 27 |
| Gastric cancer | 4 | 6 | 8 | 6 | 24 |
| Lung Cancer | 7 | 11 | 2 | 0 | 20 |
| Ovarian Cancer | 15 | 2 | 6 | 5 | 28 |
| Pancreatic Cancer | 3 | 31 | 1 | 0 | 34 |
|  | 45 | 112 | 35 | 22 | 214 |

#### Targeted CfDNA panel data

##### - Patient enrollment and sample collection

To validate the findings of this study, we enrolled 29 patients with colorectal cancer, and sequenced their cfDNA sample with a comprehensive cancer panel. The study was approved by the Ethics Committee of the First Affiliated Hospital of Nanjing Medical University. All participants involved in this study have signed informed consent forms. All cancer patients and their TNM stages were confirmed by colonoscopy and pathological examination. The blood samples from the cancer patients were collected when the patients were at the first time of diagnosis, before tumor resection or therapy. The clinical data for all participants is listed in Table 2.

**Table 2:**
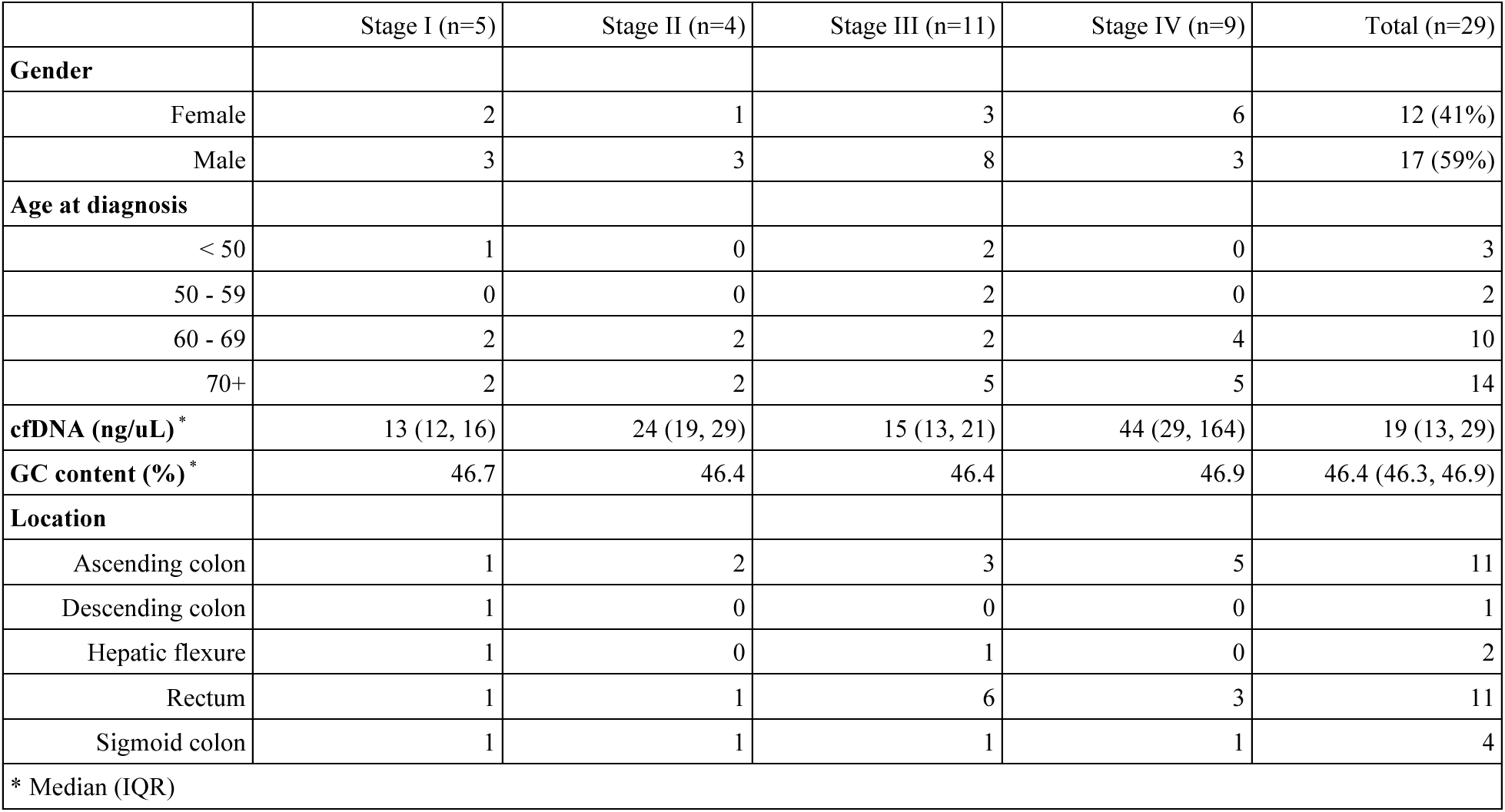
clinical characteristics of colorectal cancer patients (targeted panel data)

##### - cfDNA extraction and NGS library preparation

Blood samples, about 10 ml from each individual, were collected using the Apostle MiniMax cf-DNA Blood Collection Tube (Apostle; San Jose, CA, USA) and processed within seven days from the collection. The cfDNA extraction from 2 to 5 ml of each plasma sample was conducted using the Apostle MiniMax High Efficiency cfDNA isolation kit (Apostle; San Jose, CA, USA), with adherence to the manufacturer’s protocol with slight modifications.

The targeted sequencing procedure was applied to the 29 cancer samples using an input cfDNA range of 10-50 ng. The Agilent SureSelect Cancer All-in-one (AIO) solid tumor panel (Agilent; Sunnyvale, CA, USA) was used for the NGS library preparation. This panel contains 151 clinically validated cancer-related genes spanning a total of 2,652 genomic regions. The entirety of the genome coverage is approximately 1.32Mb, which is about 0.044% of the human genome. The median of the targeted genomic regions stands at 371bp, with a significant majority (97%) of targeted regions being under 1Kb and a near entirety (99%) under 2Kb. We removed the targeted regions if there exists one or more cancer samples with a read-depth below than 10. This removal left 2,412 targeted regions (in total 1.16Mb of the targeted region). Post-filtering, the mean read-depth for the targeted sequencing data rested between 40-50x. All sequencing procedures were executed on the Illumina HiSeq 4000 platform (Illumina; San Diego, CA, USA).

#### - Processing of targeted sequencing data and bioinformatics pipelines

A concise overview of the data processing involved first filtering the raw sequencing data of cfDNA samples using fastp (v0.23.2) to remove adapter sequences. The sequencing short-reads were then aligned to the UCSC human reference genome hg19 through the Burrows-Wheeler aligner (BWA, v0.7.17). Marking of read duplicates and the local realignments were accomplished with The Genome Analysis Toolkit (GATK 4.1.5.0), and any short-reads with a Phred quality score less than 30 were removed from the dataset. The cfDNA sequencing read fragment lengths and other downstream analysis tasks were accomplished using the R packages GenomicAlignments and GenomicRanges.

## Results

### Construction of cfDNA fragmentation profiles

The cfDNA fragmentation profile consists of a series of ratios to capture the cfDNA fragment patterns across specific genomic regions or the entire genome. Within the profile, each data point represents the ratio of short to long cfDNA fragments within a given genomic region. Cristiano and his colleagues [29] established this profile using 5Mb non-overlapping genomic segments over the whole genome. In this work, we used a similar scheme to construct the cfDNA fragmentation profile for the whole-genome cfDNA fragmentation data. We categorized the short cfDNA fragments as those with lengths from 90bp to 165bp, and the long cfDNA fragments as those ranging from 166bp to 250bp (Figure 2).

**Figure 2.**
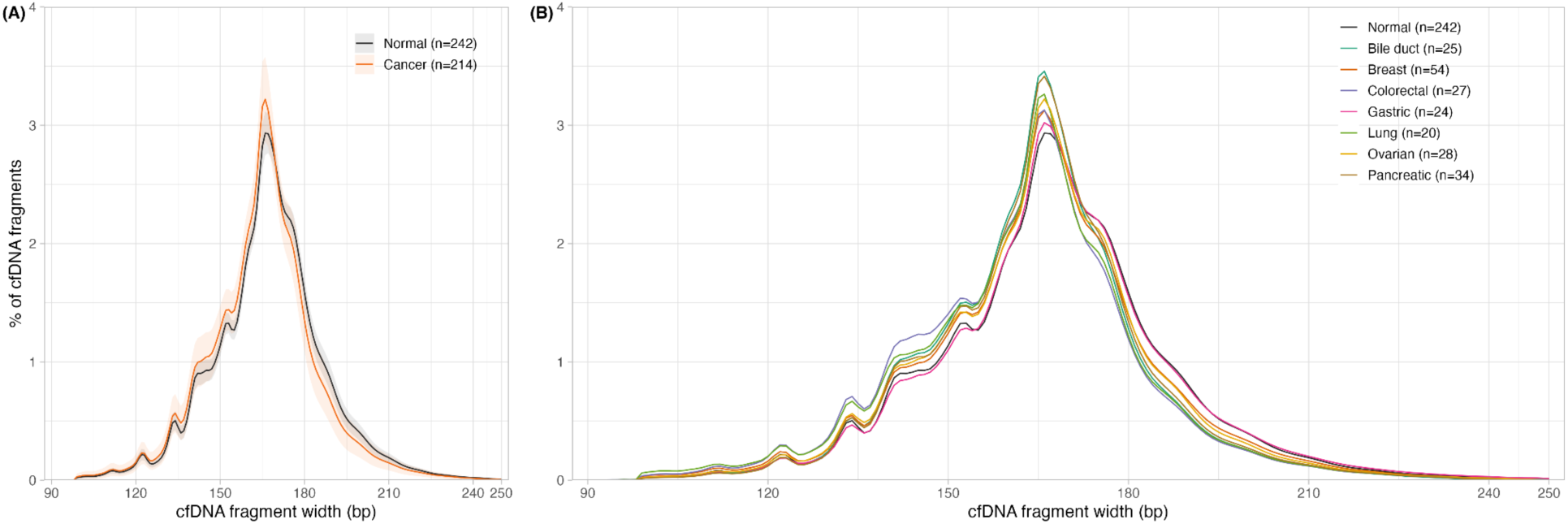
cfDNA fragment length distributions. (A) Comparison of cfDNA length distributions between healthy individuals (black, n=242) and cancer patients (red, n=214), where the ribbons denote the 95% confidence intervals. (B) Comparison of cfDNA length distributions across 7 cancer types.

For the targeted panel data, we initiated by concatenating all the targeted regions based on their genomic coordinates. Subsequently, we divided this amalgamated genomic area into non-overlapping bins, each bin at 20kb in width. Cristiano et al. suggested that a GC-content correction procedure may potentially reduce the bias introduced during the high-throughput sequencing [30–32]. However, considering the smaller bin width may lead to greater variations in the number of fragments of these bins, we skipped this phase to avoid many *NA* values introduced during the GC-correction. By comparing the profiles with and without GC-correction, No evident differences were observed for the cfDNA fragment profiles with and without GC-correction for this targeted panel data (Figure S1).

### Association analyses of cfDNA fragmentation profiles and cancer stages

We constructed cfDNA fragmentation profiles from 214 whole-genome cfDNA cancer samples across seven cancer types: bile duct, breast, colorectal, gastric, lung, ovarian, and pancreatic cancers. In Figure 3A, we employed circos plots [35] to visualize the fragmentation profiles of these cancer types according to the cancer stages I, II, and III/IV. The first circos plot depicts the median fragmentation profile consolidated from all cancer samples. The subsequent plots illustrate the median profiles for each cancer type, segmented by cancer stages. As controls, we integrated the median fragmentation profiles of healthy individuals into these plots [36]. As shown in the first circos plot, subtle distinctions are presented among the median fragmentation profiles across the three cancer stages for all cancer types included. However, the divergence was demonstrated in the fragmentation profiles of cancer stages in each cancer type at varying degrees. For instance, the fragmentation profiles of colorectal cancer cascade in an order from healthy, transitioning to stages I, stage II, and stages III/IV. In contrast, for lung cancer, the profiles of stages I and II cluster closely to each other, while the profile of stage III/IV markedly distances from these two stages.

**Figure 3.**
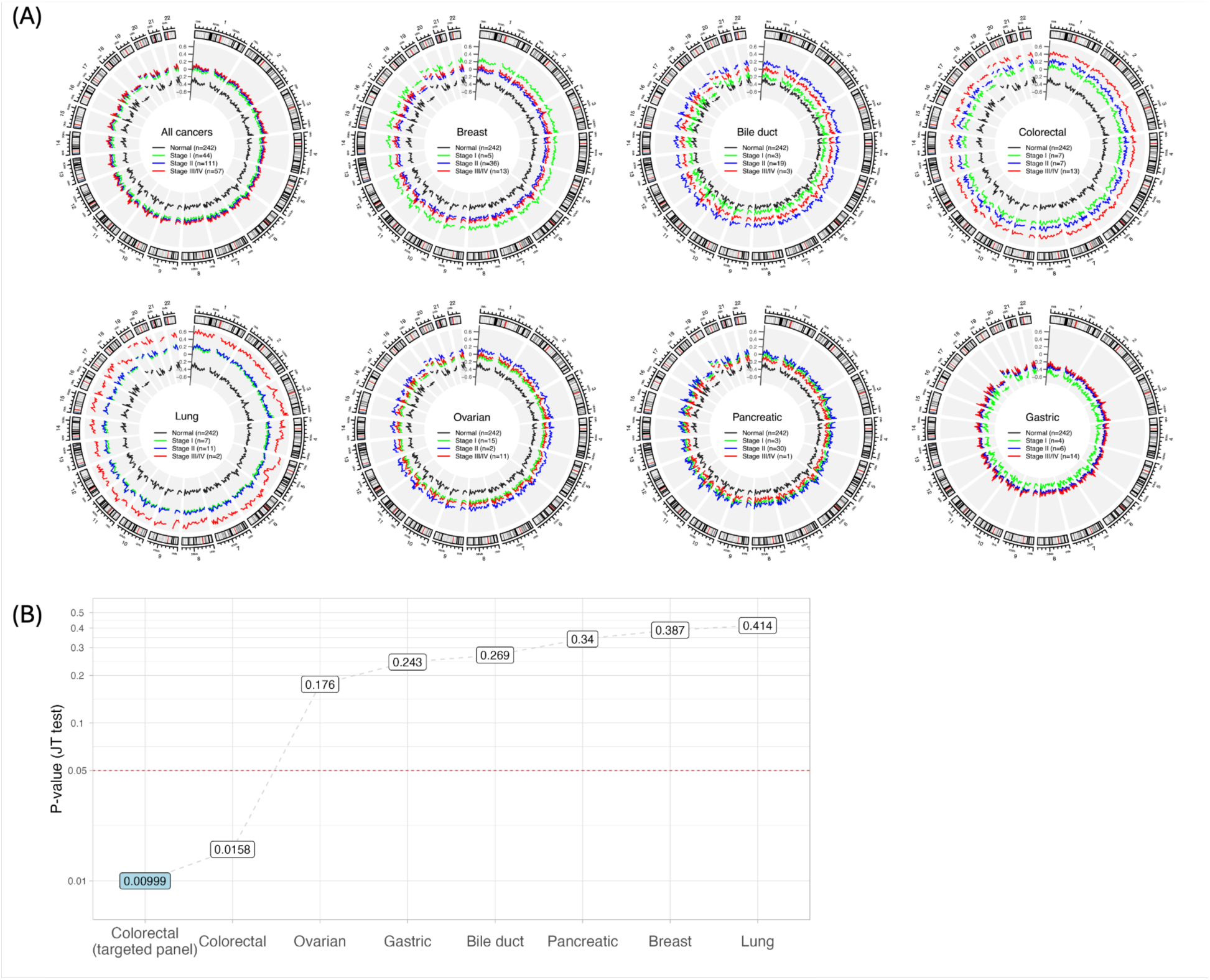
Analyses of the correlation between cfDNA fragmentation patterns and cancer stages across various cancers. (A) Circos plots illustrate genome-wide cfDNA fragmentation patterns in 5Mb intervals. For each cancer’s WGS data, median cfDNA fragmentation patterns are color-coded by stages: I (green), II (blue), and III/IV (red). Patterns from 242 healthy individuals are shown in black. The first plot represents median fragmentation patterns across all 214 cancer samples, while subsequent plots detail individual cancer types. (B) Jonckheere’s trend test determines the significance of the association between fragmentation patterns and ordered cancer stages for each type. The test is also applied to a targeted panel dataset (highlighted in light blue). Cancers are ranked based on their *p*-values; a reference line at a 0.05 cutoff is denoted in red.

The circulating tumor DNA (ctDNA) has more variability in length, and compared to cfDNA from healthy cells, the consensus of studies has indicated the average length of ctDNA is shorter in the mononucleosome range. Hence, we hypothesized that the cfDNA fragmentation profile, reflecting the ratios of short-to-long cfDNA fragments over genome regions, would elevate with the progression of disease. To test this, we applied Jonckheere’s trend test to the compiled cfDNA fragmentation profiles. Jonckheere’s trend test is a non-parametric statistical test designed to determine if there exists a significant trend across ordered groups. Using the fragmentation profiles from healthy samples as a reference, we anticipate that if the hypothesis stands, the fragmentation profiles of a specific cancer type could be in a sequential order – starting from the early cancer stage and advancing to the more advanced cancer stages. The Jonckheere’s trend test enables a quantitative assessment into the correlation between cfDNA fragmentation patterns and the progression of cancer.

Figure 3B summarizes the results of Jonckheere’s trend test across various cancer types, ranked by the *p*-values in an ascending order. Here, only the *p*-value of colorectal cancer (*p*-value = 0.0158) falls below the conventional statistical significance threshold of 0.05, suggesting a statistically significant association between cfDNA fragmentation profiles and the progression of cancer stages.

### Validating the association of cfDNA fragment profiles and cancer stages in CRC

To confirm the association between cfDNA fragmentation profiles and the stages of colorectal cancer, we sequenced for 29 patients with colorectal cancer using a comprehensive cancer gene panel (read depth 40-50x). This panel targets 151 clinically validated cancer-associated genes (Table S1), covering roughly 1.32 MB or about 0.044% of the entire human genome.

Figure 4A denotes the cfDNA fragment length distributions of the WGS and targeted panel data. Although the targeted panel covers a small fraction of the human genome, the overall fragment length distributions between the two datasets are congruent. In addition, both datasets display the oscillated waves occurring every 10bp in the 100bp to 150bp range, a typical trait of cfDNA fragment length distribution, indicating the targeted panel data can essentially reflect the characteristics of cfDNA fragmentation patterns.

**Figure 4.**
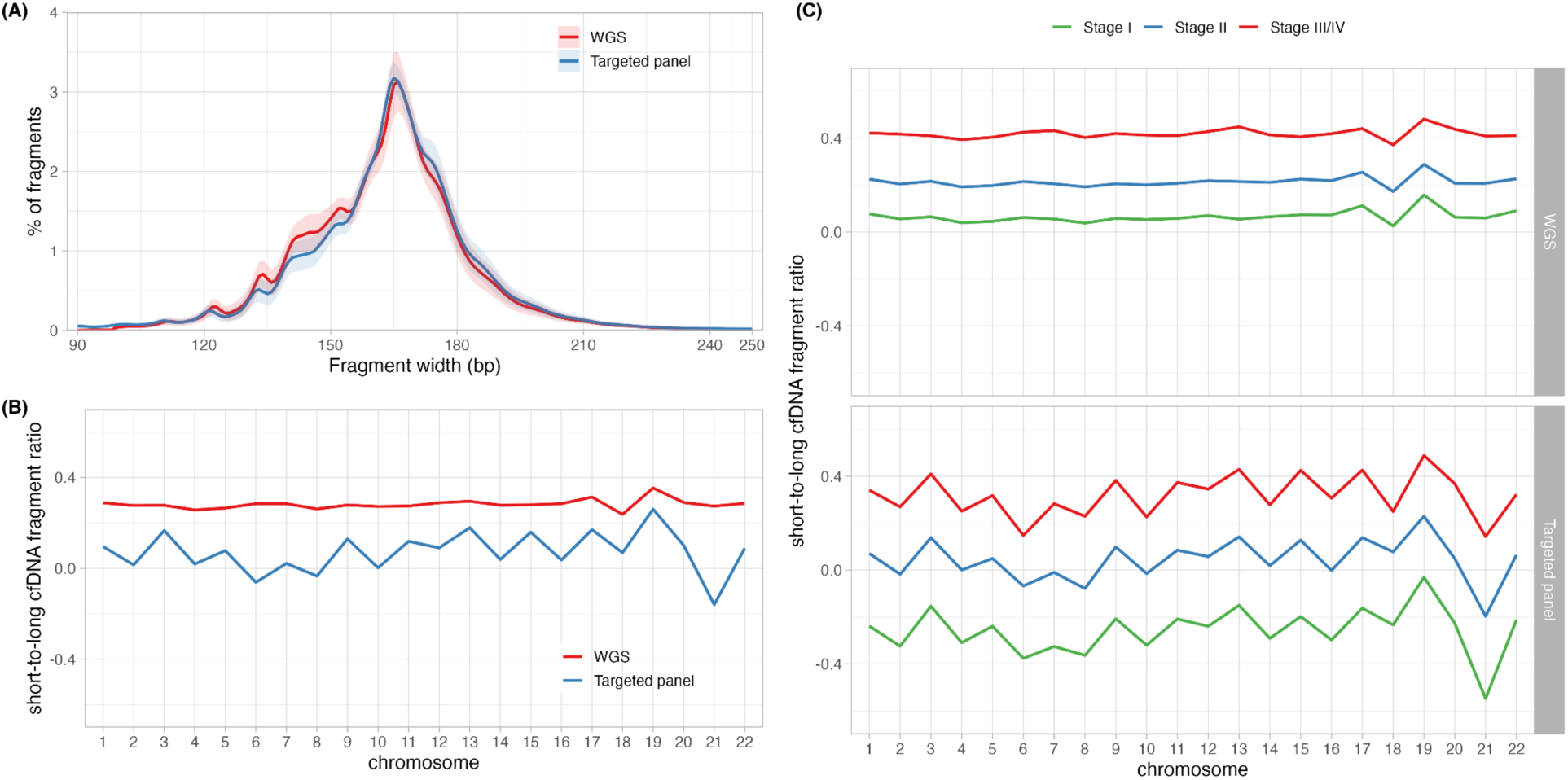
Comparison of WGS and targeted panel datasets of colorectal cancer. (A) The cfDNA fragment length distributions of both datasets align closely. Oscillatory patterns appear every 10bp in the range of 100bp to 150bp, with fragment length with a peak at 165bp. (B) Median cfDNA fragmentation profiles from both datasets are compared at the chromosomal level. (C) Fragmentation profiles from the two datasets, categorized by cancer stage, demonstrate similar progression trends. Despite differences in genome coverage and sequencing techniques between two datasets, their cfDNA fragmentation patterns consistently align with cancer stages.

To compare the cfDNA fragmentation profiles of two datasets, we constructed profiles for both datasets at the chromosome level (chr1 to chr22). Fig 4B and Fig 4C demonstrate the overall median profiles of these two datasets according to cancer stages. Although owing to different genomic coverage, little similarity is shown between the overall fragmentation profiles of these two datasets (Fig 4B), when we compare them regarding their cancer stages, these profiles are stratified in the same order with respect to cancer stages. The Jonckheere’s trend test, applying to the fragmentation profiles of the targeted panel data at a bin width of 20Kb, achieved a *p*-value of 0.01 (Fig 3B). Above all, we conclude that for colorectal cancer, the cfDNA fragmentation profiles consistently align with cancer stages in order, and hence the cfDNA fragmentation profiles of CRC could be used as a biomarker to distinguish cancer stages.

### Frag2Stage: using cfDNA fragmentation profiles to classify cancer stages of CRC

Our insights from the previous findings inspired the development of “frag2stage”, a machine-learning model that leverages cfDNA fragmentation profiles to classify CRC cancer stages. Frag2stage utilizes a *l1*-regularized linear regression model (LASSO) to predict cancer stages from cfDNA fragmentation profiles. We employed a 5-fold cross-validation with 10 repeats to avoid overfitting. For imbalanced datasets, we adopted the up-sampling strategy, and repeated this step 50 times to offset the potential biases introduced from sampling. Our method was implemented in R (version 4.2.1). We used *caret* package (version 6.0.94) to control cross-validation and model training and used *glmnet* package (version 4.1.8) for LASSO.

For the whole-genome cfDNA data, our analysis encompassed two classification tasks: differentiating stage I from stage IV samples and distinguishing between stages I/II and stages III/IV samples. For the targeted panel data, we introduced an additional layer to discern stages I/II from stage IV samples. The whole-genome data was analyzed at a bin width of 5MB (resulting in 518 bins); while the targeted panel was assessed at a bin width of 20KB (56 bins). Table 3 summarizes the prediction performance with three metrics: AUC (Area Under Curve) of ROC (Receiver operating characteristic), precision 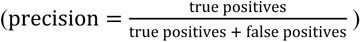 and F1-score 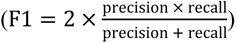. Figure 5A illustrates the classification outcomes using the ROC curves. In detail, the frag2stage model, when applied to the whole-genome data, adeptly differentiates between stages I/II to III/IV and stages I to IV, with the AUC values of 0.73 and 0.77 respectively. For the targeted panel data, the resulting AUC values stood at 0.68, 0.79, and 0.99 for the classifications of stages I/II to III/IV, stages I/II to IV, and stages I to IV, respectively. For simplicity, we incorporate ROC-AUC values in the downstream analysis.

**Figure 5.**
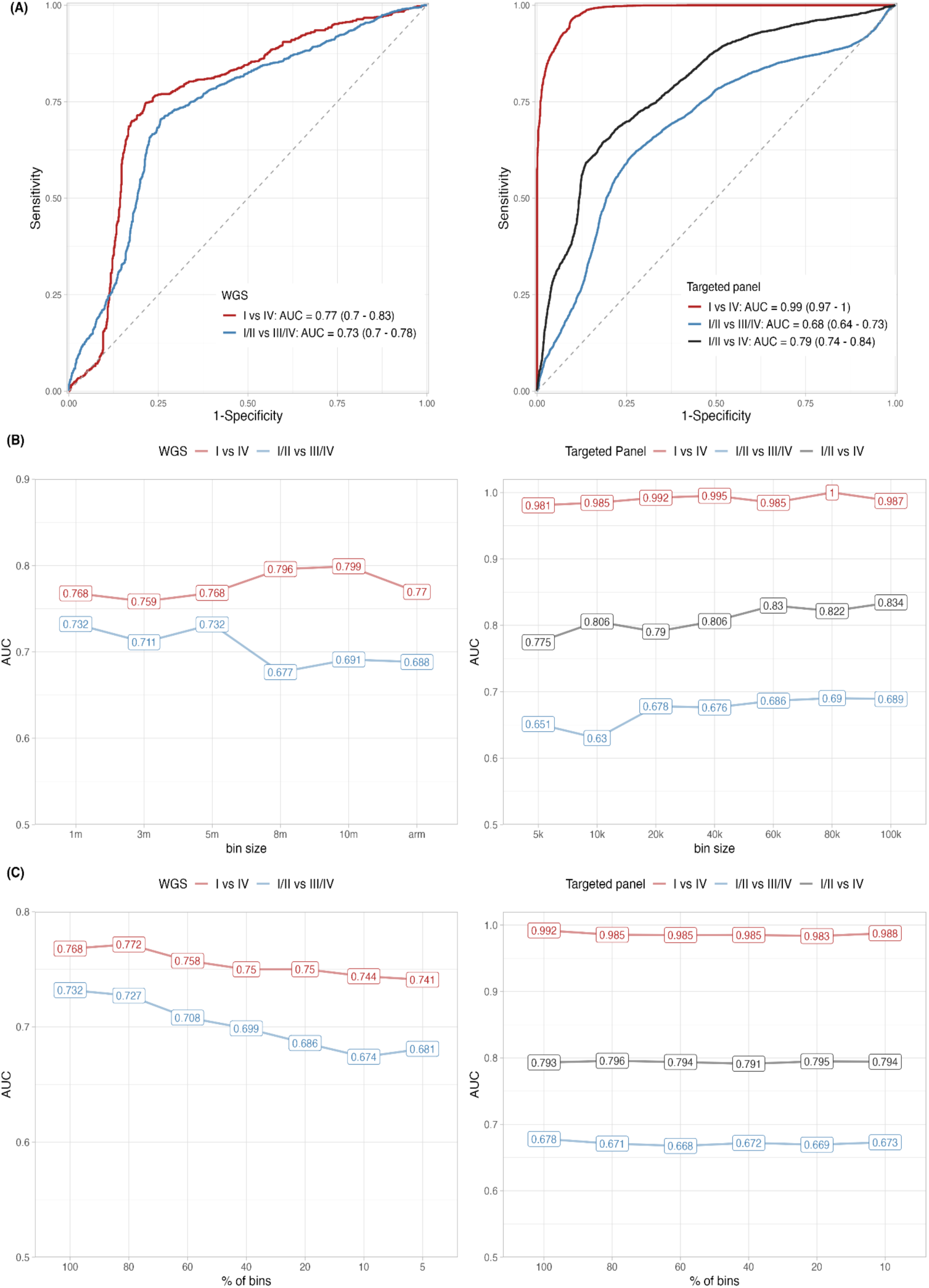
Cancer stage prediction using frag2stage across two datasets. (A) ROC curves display the efficacy of frag2stage in distinguishing cancer stages based on cfDNA fragmentation profiles. For the WGS dataset, AUC values are 0.73 (stage I/II vs. III/IV) and 0.77 (stage I vs. IV), respectively. For the targeted panel dataset, AUCs are 0.68 (stage I/II vs. III/IV), 0.79 (stage I/II vs. IV), and 0.99 (stage I vs. IV), respectively. (B) Evaluating the robustness of frag2stage subject to bin width variation. The plots illustrate ROC values based on varying bin widths for both datasets. (C) Assessing the robustness of frag2stage against genome coverage variability. When applied to cfDNA fragmentation profiles with randomly selected bins at different degrees, the prediction performance of the WGS data drops gradually, while the targeted panel data retains consistent performance.

**Table 3:**
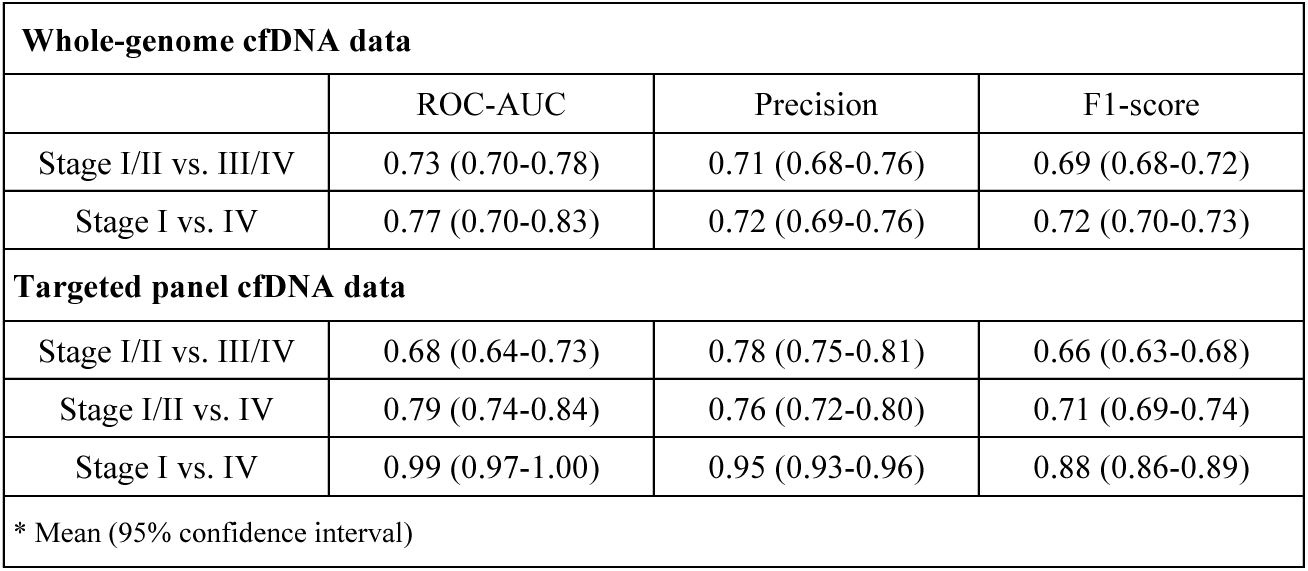
Performance of frag2stage method on the whole-genome and targeted panel datasets.

### Evaluation of the robustness of frag2stage

Frag2stage is based upon cfDNA fragmentation profiles, and therefore we assessed the robustness of the model against two variables, the bin width of fragmentation profiles and the coverage of genomic regions that are used in the classification.

To understand how the performance of our model varies with respect to genomic bin width, we applied frag2stage across a spectrum of widths. As illustrated in Figure, for both whole-genome and targeted panel datasets, the shift of prediction performance remains modest, hovering within a 0.06 margin in the AUC values.

Next, we tested how frag2stage would perform when only a fraction of data points in the cfDNA fragmentation profile are employed. This involved random selection of varying percentages of data points from the profiles, upon which frag2stage was applied. To reduce evaluation biases, this randomized sampling was repeated 50 times at each percentage (Fig 5C). For the whole-genome data, the AUC values across all two stage classifications drop gradually within 0.05. In contrast, the AUC values of the targeted panel data for all classifications remains consistent with minor fluctuations.

In all, these analyses reveal the cfDNA fragmentation patterns are a reliable biomarker for distinguishing stages of colorectal cancer. Remarkably, even with minimal genomic data slices, frag2stage can classify cancer stages at a satisfactory precision.

## Discussion

In this study, we explored the correlation between cfDNA fragmentation patterns and cancer TNM stages across seven cancer types. We discovered that the relationship between these patterns and cancer stages varies depending on cancer type. Tumor heterogeneity [33] and dynamic nature of cancer [34] could be the primary factors that contributed to this variability. Specifically, the inherent heterogeneity within tumors creates multiple subcellular populations, and each endowed with distinct genetic and epigenetic characteristics, leading to various cfDNA fragmentation patterns in different cancer types. Furthermore, as tumors progress, alterations in genetic composition, metabolism and other biological mechanisms may evolve in distinct directions in different cancer origins, leading to diversity in this relationship. Additionally, the empirical nature of the TNM staging system might not invariably correlate with genomic alterations, introducing an additional layer of complexity. Notably, we identified a strong association between cfDNA fragmentation patterns and cancer stages in colorectal cancer, and this relationship is further confirmed by an independent targeted panel data.

Further, we developed a machine learning model to classify cancer stages for colorectal cancer. We validated our model on two independent datasets and achieved promising AUC values. One intriguing result from our study is that our model can distinguish cancer stages using cfDNA fragmentation information from only a small portion of the genome, suggesting that alterations in cfDNA fragmentation patterns associated with the CRC progression is a university phenomenon over the whole human genome.

With the advance of high-throughput sequencing technologies, the cost of NGS-based diagnostic tests has significantly decreased, making them commonplace in certain clinical settings. In this study, we demonstrated that cfDNA fragmentation information, derived from targeted sequencing (less than 50 US dollars), can distinguish the TNM stages of colorectal cancers. While we were not able to access the DNA genomic composition of the whole-genome data, the cfDNA fragmentation profiles still allowed us to differentiate between early and advanced stages. We believe the performance of our method can further be improved by integrating other genetic data and epigenetic data.

To our knowledge, this is the first study to exclusively use cfDNA fragmentation data for cancer prognosis predictions. Our results hint at a potential non-invasive method for determining cancer stages, especially for colorectal cancer. As a proof-of-concept study, our findings offer a promising direction for liquid biopsy in cancer diagnosis and personalized medicine, setting the stage for future research and potential clinical applications.

## Data and code availability

Sequencing data and codes used in this study will be available to the public on acceptance.

## Data Availability

All data produced in the present study are available upon reasonable request to the authors.

---

**Figure S1.**
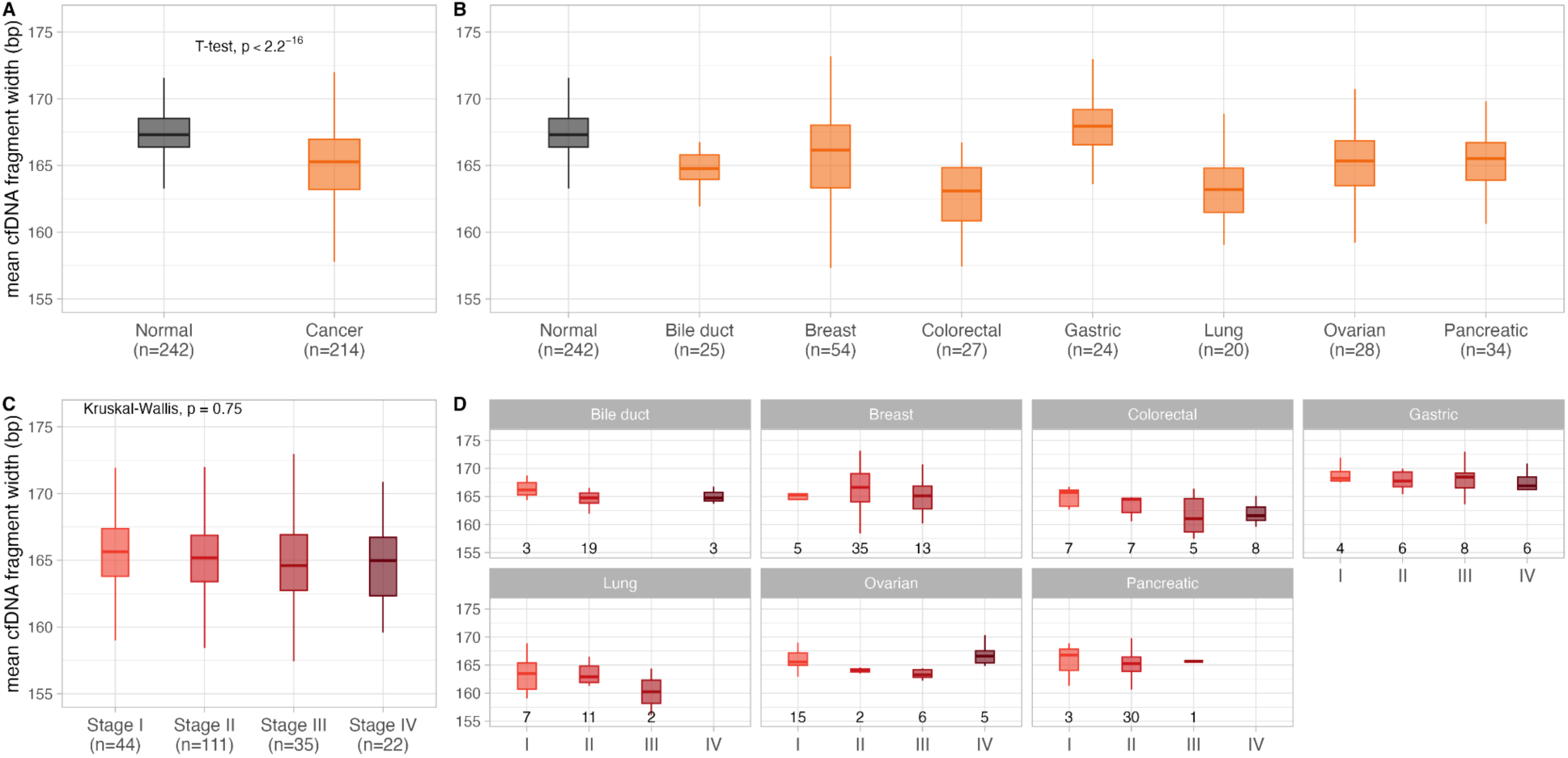
Analyses of mean cfDNA fragment width. (A) Comparison of mean cfDNA fragment widths between healthy individuals (n=242) and cancer patients (n=214). (B) Comparison of mean cfDNA fragment widths of healthy individuals and seven cancer types. (C) Comparison of mean cfDNA fragment widths across cancer stages. (D) For each cancer type, comparison of mean cfDNA fragment widths across cancer stages.

**Figure S2.**
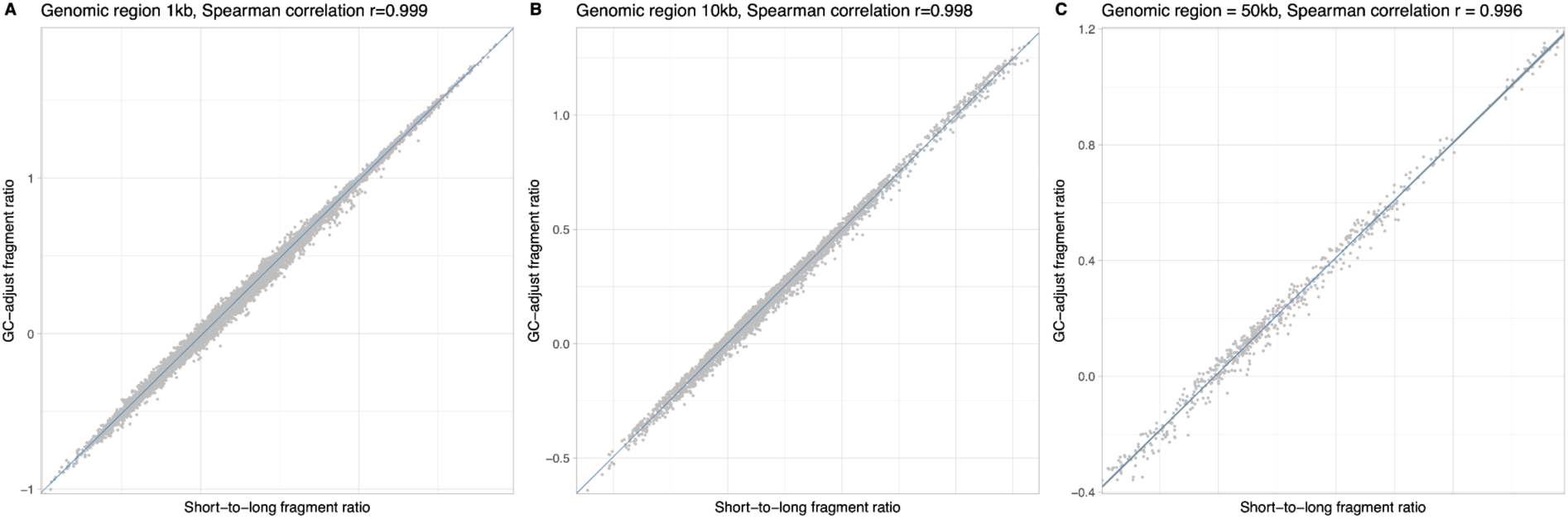
Consistent correlation of cfDNA short-to-long fragment ratios pre- and post-GC-content Adjustment. (A-C) The correlation at genomic bin sizes of 1Kb, 10Kb, and 50Kb, respectively. Each point on the plots denotes a specific data entry in a cfDNA fragmentation profile. The *x*-axis represents the short-to-long fragment ratio prior to GC-content adjustment, while the *y*-axis represents the ratio post-adjustment.

**Supplementary table 1:**
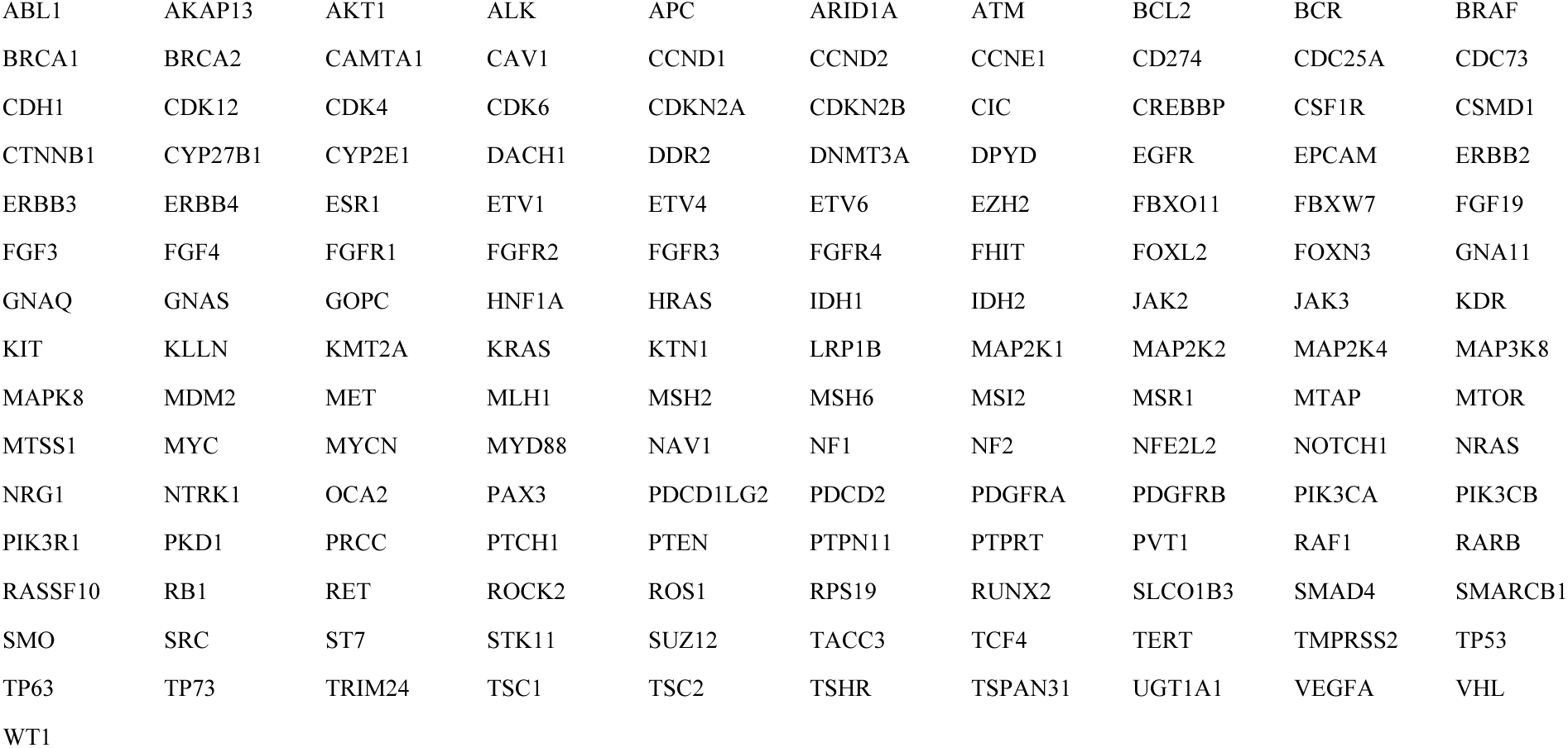
list of the 151 targeted genes of the AIO solid-tumor panel.

